# Real-World Effectiveness of Deltamethrin-Clothianidin (FLUDORA FUSION) in Indoor Residual Spraying in Sub-Saharan Africa: A Systematic Review Protocol

**DOI:** 10.1101/2024.06.01.24308309

**Authors:** Moses Ocan, Nakalembe Loyce, Kevin Ouma Ojiambo, Geofrey Kinalwa, Alison A. Kinengyere, Sam Nsobya, Emmanuel Arinaitwe, Henry Mawejje

**Author notes:** **Correspondence:** Dr. Moses Ocan, Africa Center for Systematic Reviews and Knowledge Translation, Makerere University College of Health Sciences & Department of Pharmacology, School of Biomedical Sciences, College of Health Sciences, Makerere University, P.O Box 7072, Kampala, Uganda. These authors contributed equally to this work (Nakalembe Loyce, Kevin Ouma Ojiambo). **Competing interests:** None declared. **Data Availability:** The data from this study will be publicly available from the published studies included in the review.

## Abstract

**Background:** Indoor residual spraying (IRS) is a core insecticide-based vector control tool employed in most malaria-affected settings globally. However, mosquito vectors have developed resistance to nearly all the insecticides currently used in IRS. This has necessitated a transition to new classes of insecticides from mostly using Dichlorodiphenyltrichloroethane (DDT) and pyrethroids from 1997 to 2010, to carbamates from 2011 and organophosphates from 2013. In addition, other vector control measures like the use of long-lasting insecticide-treated bed nets (LLINs) have also been employed for malaria control. Despite the implementation of these mosquito vector control interventions, malaria remains a disease of public health concern especially in sub-Saharan Africa which bears over 90% of the disease burden. This review will thus collate evidence on the effectiveness of IRS for malaria control in sub-Saharan Africa.

**Methods and analysis:** The systematic review will be done following *a priori* criteria developed using PRISMA guidelines. Articles will be obtained through a search of Medline via PubMed, Scopus, and Embase databases. The Mesh terms and Boolean operators (“AND,” “OR”) will be used in the article search. Additionally, websites of malaria research institutions will be searched. Article search will be done by two independent librarians (AAK and RS). All identified articles will be transferred to Epi-reviewer v6.15.1.0 software. Article screening and data abstraction will be done in duplicate by four reviewers (KO, LN, GK and MO) and any further disagreements will be resolved through discussion and consensus. Data analysis will be done using STATA *v*17.0. Heterogeneity in the articles will be assessed using the I^2^ statistic. Publication bias will be assessed using a funnel plot.

**Results:** The findings of this review will help generate evidence on the effectiveness of indoor residual spraying using WHO pre-qualified insecticides in malaria control in sub-Saharan Africa.

This protocol was registered in PROSPERO (https://www.crd.york.ac.uk/prospero/) registration number CRD42024517119

## INTRODUCTION

Indoor residual spraying (IRS) is one of the current WHO-recommended insecticide-based vector control strategies which is intended to reduce and, ultimately, interrupt malaria transmission (1). According to WHO, IRS is the application of a long-lasting, residual insecticide to potential malaria vector resting surfaces such as internal walls, eaves and ceilings of all houses or structures (1). Indoor residual spraying aims at i) reducing the vector’s lifespan to less than the time it takes for the malaria sporozoites to develop, ii) reducing vector density by immediate killing, and iii) reducing human–vector contact through a repellent effect, thereby reducing the number of mosquitoes that enter sprayed rooms (1). In 2010 and 2013, approximately 185 and 124 million people were protected by the IRS representing 6% and 4% of the global population at risk respectively (1). In the WHO African region, the number of people protected by the IRS increased from 10 million in 2005 to 78 million in 2010 (1).

Some of the commonly recommended classes of insecticides used as active ingredients for IRS include pyrethroids (such as alpha-cypermethrin, deltamethrin, lambda-cyhalothrin), organophosphates (e.g. malathion, pirimiphos-methyl), carbamates (i.e. bendiocarb and propoxur), neonicotinoids such as clothianidin and organochlorines e.g. (dichlorodiphenyltrichloroethane, DDT) (2). These insecticide classes have different residual activity, cost and efficacy in the field (3). Among these, pyrethroids are the most frequently used insecticides due to their relatively low toxicity to humans, fast knockdown effect and cost-effectiveness (2, 4, 5).

The IRS is however affected by the high costs, complex implementation logistics, and community acceptance (6). Additionally, the emergence of resistance to most insecticides used for IRS has also been reported in most sub-Saharan African countries (7, 8). Resistance of malaria vectors to pyrethroids and other classes of insecticides is widespread in sub-Saharan Africa (5, 7, 8). Resistance to IRS is driven by selection pressure placed on resistance genes, heavy reliance on one class of insecticides for vector control and continued use of the same chemical classes as agricultural pesticides (2).

Several strategies have been used to delay the development of resistance to the already existing insecticides including; regularly changing insecticides (rotations), use of a combination of insecticides with different modes of action, deployment of insecticides of different modes of action in neighbouring geographical areas (mosaic spraying) and co-deployment of different interventions in the same place (8, 9). Combination insecticide mixtures have the dual potential to improve malaria vector control in addition to managing resistance (10). One of the recently WHO-approved insecticide mixtures is a formulation of a wettable powder product containing 500 g/kg of clothianidin and 62.5 g/kg of deltamethrin (Fludora® Fusion) (10).

Despite the widespread application of indoor residual spraying and other mosquito vector control measures, malaria remains a disease of public concern especially in sub-Sahara Africa. This is further worsened by emerging and widespread resistance to insecticides used in IRS and bed nets. The introduction of interventions such as combination insecticide preparations could potentially improve the effectiveness of insecticide-based mosquito vector control interventions including IRS. Individual studies have assessed the efficacy of insecticides used in IRS in most malaria-affected regions. With malaria prevalence remaining high in sub-Saharan Africa there is a need to establish the real-world effectiveness of recently approved combination and single insecticide preparations used in IRS. However, this remains unknown in most high malaria endemic settings of sub-Saharan Africa which affects decision-making on insecticide rotation strategies for mitigating resistance development. This review will collate context-specific evidence on the efficacy of WHO pre-qualified combination and single-agent insecticide preparations in indoor residual spraying for malaria control in sub-Sahara Africa. This will help inform policy and decision-making on insecticide rotation strategies in the fight against widespread insecticide resistance development.

### Rationale

Recently, WHO pre-qualified insecticides for IRS including FLUDORA FUSION (Deltamethrin-Clothianidin), Clothianidin, and Broflanilide. Together with other insecticides their use in insecticide-based interventions has contributed to approximately 78% of the reduction in malaria burden in sub-Saharan Africa since 2000 (11). However, the implementation of the IRS is currently faced with the challenge of the emergence and spread of insecticide resistance (11). The emergence of insecticide resistance in *Anopheles* mosquitoes in sub-Saharan Africa has implications for vector control interventions. This has led to the transition to different classes of insecticides used in IRS. Several reviews have recently synthesized evidence on the effectiveness of IRS on malaria transmission however, these have several limitations ((12–14). Most of these reviews focused on measuring effectiveness with the majority assessing only the effect of IRS in reducing the malaria burden (12–14). A review by Giming et al., (2023) focused only on reactive IRS application in the control of malaria (13). However, malaria remains prevalent in most countries in sub-Saharan Africa which commonly use proactive IRS application (13). Another systematic review by Pryce et al, (2022) focused on the effect on malaria of additionally implementing IRS, using non-pyrethroid-like or pyrethroid-like insecticides, in communities currently using ITNs, hence results may not directly provide evidence on the actual effect of IRS when used alone (14). In a review by Zhou et al., (2022), pyrethroids were identified to show the greatest performance in malaria control while Pryce et al., 2022 showed that adding non-pyrethroid insecticide to bed nets had the greatest effect on malaria burden. The review by Pryce et al., (2022) did not find a significant effect of adding pyrethroid insecticide to bed nets in malaria control. Additionally, a review by Zhou et al., 2022 included only studies that used DDT, methylcarbamate and primiparous-methyl insecticides only (12). There is thus still an evidence gap on the effectiveness of other WHO pre-qualified insecticides for IRS including combination products. Additionally, the WHO recently approved combination/mixed insecticide preparations for use in IRS such as Fludora fusion a combination of Clothianidin-Deltamethrin and single insecticide preparations including Broflanilide and Clothianidin. However, no review has evaluated the available evidence of their impact on the malaria burden in sub-Saharan Africa. The current review therefore seeks to collate evidence on the efficacy of recent WHO pre-qualified single and combination insecticide preparations used in IRS for malaria control in sub-Saharan Africa. The review will focus on the IRS effect on mosquito vector-related outcomes such as density, mortality/susceptibility, knockdown rate, resistance genes and malaria indices in communities.

### How the intervention might work

Indoor residual spraying is one of the preventive measures aimed at eliminating malaria globally. It involves the use of insecticides which reduce the number and longevity of mosquito vectors, thereby decreasing malaria transmission (13). Compared to long-lasting insecticide nets which provide a barrier for mosquito vectors, IRS acts in multiple ways including repellant, reducing malaria transmission and vector mortality. Furthermore, the current introduction of combination insecticide preparation improved the efficacy and potentially reduced the risk of widespread resistance development. Resistance to insecticides remains the greatest risk to IRS especially in high burden settings common in sub-Sahara Africa. The introduction of new insecticide classes in addition to combination agents could potentially be responsible for the recent gains in malaria control.

### Primary review question

1. What is the real-world effectiveness of Deltamethrin-Clothianidin (FLUDORA FUSION) insecticide used in indoor residual spraying in sub-Sahara Africa?

### Secondary review question (s)

1. What is the prevalence of mosquito vector genotypic resistance to combination and single-agent insecticides in sub-Sahara Africa?
2. What is the prevalence of kdr resistance genes among mosquito vectors in settings using combination and single-agent insecticides in sub-Sahara Africa?
3. What are the species of malaria mosquito vectors in settings using combination and single-agent insecticides in sub-Saharan Africa?
4. What factors are the factors associated with the real-world effectiveness of combination and single-agent insecticides used in indoor residual spraying in sub-Sahara Africa?

## MATERIALS & METHODS

### Protocol Registration

We shall follow Preferred Reporting Items for Systematic Reviews and Meta-Analysis Guidelines (15) to perform this systematic review. This protocol is written according to Preferred Reporting Items for Systematic Review and Meta-Analysis Protocols (PRISMA-(16) and was registered in PROSPERO (https://www.crd.york.ac.uk/prospero/) with registration number CRD42024517119

### Eligibility criteria Inclusion

- Articles that report the impact of IRS using WHO pre-qualified insecticides.
- Articles published in peer-reviewed journals.
- Articles published in all languages (no language restriction)

### Exclusion

- Articles that do not segregate the effect of multiple mosquito vector control measures on malaria burden in sub-Saharan African countries.
- Articles that report on vector insecticide susceptibility outside the context of the IRS
- Articles not reporting ethical review and approval.

### Identification of Primary Studies

An experienced librarian (AAK) and the principal investigator (OM) will independently search for the articles from established databases. The articles from two independent searches will then be merged in EndNote software and duplicates removed.

### Information Sources

Articles published in peer-reviewed journals reporting on the effectiveness of indoor residual spraying (IRS) vector intervention for malaria control will be searched from; Google Scholar, and MEDLINE via PubMed, Scopus, and Embase databases. The search will cover a period from 1990 to date and will include sub-Saharan African countries. Furthermore, we will conduct a hand search on institutional websites for any relevant grey literature. We will also screen through reference lists of included studies for additional eligible articles.

### Search strategy

The scoping literature search was finished on January 18, 2024 however, the full article search has not yet been carried out. To find relevant articles based on PICOST, the search terms listed below will be used in the full article search. Boolean operator “OR” will be used to combine terms that relate to the same PICOST element, while “AND” will be used to join terms that relate to separate concepts or PICO categories.

### Search terms

The following search terms will be used; ‘Fludora Fusion’, ‘deltamethrin’, ‘bendiocarb’, ‘primiphos-methyl’, ‘DDT’, ‘dichlorodiphenyl-trichloroethane’, ‘malathion’, ‘temephos;, ‘fenitrothion’, ‘cypermethrin’, ‘chlothianidin’, ‘insecticide’, ‘Actellic’, ‘chlorfenapyr’, ‘propoxur’, ‘pyrethroid’, neonicotinoid’, ‘Sumishield’, ‘*Anopheles gambiae s.l’, ‘Anopheles funestus’, ‘Anopheles arabiensis’, Anopheles stephensi’, ‘Anopheles pharoensis’, ‘mosquito’, ‘malaria vector mosquito’, ‘mosquitoes’, ‘susceptibility’, ‘efficacy’, ‘sensitivity’, ‘knock-down’, ‘mortality’, ‘delayed mortality’, ‘residuality’, ‘residual life’, ‘Indoor residual spraying’, ‘IRS’, ‘spray technique’, ‘malaria transmission’, ‘season’, ‘rainfall season’, ‘rebound malaria epidemics’, ‘malaria epidemics’, ‘insecticide resistance’, ‘insecticide tolerance’, ‘resistance’, ‘resistance genes’, ‘molecular marker’, ‘resistance alleles’, ‘sub-Sahara Africa’.* The search string will be developed using the above terms.

### Data Management and Study Selection

For the initial management of references from search results, EndNote *v*20 software will be used. The articles will then be exported to Epi-reviewer v6.15.1.0 software. The articles will then be screened in duplicate using predetermined eligibility criteria. The screening will be done independently in duplicate by the review team (MO, GK, KOO and LN) in EPPI-Reviewer v6.15.1.0, using a screening tool developed *a priori* and piloted using 10% of the search yield. Kappa agreement of 80% will be used and any disagreements between the reviewers resolved through discussion and consensus. Any further disagreements will be referred to the tiebreaker (OM).

### Data Abstraction and Coding

The data abstraction tool will be created in Microsoft Excel spreadsheet 2007 and piloted using 10% of the eligible studies. The final tool will then be uploaded in EPPI-Reviewer v6.15.1.0. The coding process will be carried out independently in pairs by research team members (MO, GK, KOO and LN). Kappa agreement of 80% will be used and any disagreements resolved through discussion and consensus. The data will later be validated for quality control by an independent senior reviewer (OM) to ensure completeness and correctness.

### Data Items

The following data categories will be abstracted: administrative information (author, year of publication, DOI, country/region, funding source), methods (study design, population, sample size), and results (insecticide efficacy, resistance, knockdown effect, susceptibility) (Table 2).

**Table 1:** Key elements of the systematic review question.

| Element of review question | Description |
| --- | --- |
| P: Population | All people living in malaria-affected settings of sub-Saharan Africa.<br>All malaria mosquito vectors in malaria-affected settings in sub-Saharan Africa. |
| I/E: Exposure | Application of combination insecticide Deltamethrin-Clothianidin in indoor residual spraying |
| C: Comparator | None |
| O: Outcome | <u>Primary outcome</u><br>Real-world effectiveness of Deltamethrin-Clothianidin in indoor residual spraying<br><u>Secondary outcomes</u><br>Mosquito vector knock-down rate<br>Prevalence of kdr gene<br>Mosquito vector mortality rate<br>Type and class of insecticide used in IRS.<br>Residuality of the insecticide on different wall surfaces |
|  | • Mosquito vector insecticide susceptibility (phenotypic resistance)<br>• Mosquito vector insecticide resistance genes (molecular resistance)<br>• Mosquito vector (s) (species)<br>• Community acceptability and feasibility<br>• Nature of indoor residual spraying (proactive, reactive, and focal IRS) |
| Design | • Randomized control trials (RCTs), case-control studies, Cohort studies, Interrupted time series, Before-and-after design, and cross-section studies/surveys. |
| Time | • Studies done from 1990 to date |

**Table 2:** Review results items/areas.

| Item | Description |
| --- | --- |
| 1. Administrative data | This will collect data to identify the articles including author, citation, funding, and country |
| 2. Method | Data will be collected on; study design, insecticide (name, formulation, and strength), combination, season, susceptibility procedure used, vector control interventions, population |
| 3. Results | Malaria incidence, malaria prevalence, insecticide efficacy, susceptibility (phenotypic resistance), genotypic resistance, vector mortality and knockdown effect |
| 4. Setting | Countries in sub-Saharan Africa |

### Outcomes and Prioritization

### Dependent variable

- Prevalence of malaria in communities following indoor residual spraying (IRS) using Deltamethrin-Clothianidin, and other insecticides for mosquito vector control in sub-Sahara Africa.

### Independent variables

- Mosquito vectors knockdown effect.
- Residuality (residual efficacy).
- Types of mosquito vectors
- Mosquito vector insecticide molecular resistance genes
- Insecticides used in indoor residual spraying for mosquito vector control in sub-Sahara Africa.
- Type of insecticide (single or combination compound).
- Factors associated with the efficacy of insecticides in indoor residual spraying in SSA.

### Risk of bias assessment

Two research team members (MO, GK, LN and KOO) will independently assess the methodological quality of included observational studies using a modified version of the Newcastle-Ottawa Scale (NOS) (17). The tool includes seven domains rated from 0 (high risk of bias) to 3 (low risk of bias); the mean of the domains is considered to result in a score between 0 and 3, with a higher score indicating a lower risk of bias. Consensus on any disagreements in the quality assessment will be reached after discussion and consultation with an independent senior reviewer.

For randomized controlled trials (RCTs) and non-randomized trials of interventions, we will use the Cochrane Risk of Bias tool and Risk of Bias in Non-Randomized Studies - of Interventions (ROBINS-I) tool to assess the potential risk of bias (18–20). Bias is measured as a rating (high, low, or unclear) for individual elements from five domains (selection bias, attrition bias, performance bias, reporting bias, detection bias, and other biases such as conflict of interest). In addition, the tools provide for the assessment of concerns for the applicability of the study to the systematic review which will also be assessed. The Kappa agreement of 80% will be used and any disagreement resolved through discussion and consensus. Any further unresolved disagreements will be referred to the tiebreaker (MO).

### Publication bias

The included articles will be evaluated for publication bias using the asymmetry of funnel plots and Egger’s test, as appropriate (21, 22). These rank-based data augmentation techniques are reliable for detecting publication bias caused by missing data/studies. We will create funnel plots and use their symmetry to determine the likelihood of publication bias among the articles included in the review. In the absence of missing studies, the scatter plot looks like a symmetrical inverted funnel with a wide base and a narrow top. The presence of large “holes” or asymmetry in the plot suggests publication bias, but this could also be explained by other factors such as study heterogeneity.

### Assessment of strength and confidence of cumulative evidence

A modified GRADE approach will be used to evaluate the overall strength of evidence. We will assign certainty of evidence ratings for the aforementioned outcome variables using a method developed by the GRADE Working Group (23) and we will do this in duplicate. Any disagreements will be resolved through consensus.

### Heterogeneity

The I^2^ statistic will be used to determine the degree of statistical heterogeneity in the included articles. The I^2^ statistic will display the percentage (%) of heterogeneity due to between-study variation (24–26). Heterogeneity will be classified as low (I^2^=25%), moderate (I^2^=50%), or high (I^2^>75%). Subgroup analysis will be performed on articles with low to moderate heterogeneity (27).

### Criteria for determination of independent findings

Dependency may arise at the study or intra-study levels. At the study level, the most complete and latest report, where available, will be selected in case of multiple reports of a single study. However, an integrative method will be used to treat the data from all of these reports as a single case if they cover various sub-groups or outcomes (28). Each meta-analysis will contain only one effect at the intra-study level from every investigation.

Studies that report more than one effect for distinct outcome types will be synthesized independently. Before including “synthetic effects” in a meta-analysis, we will use them to create a sample-weighted average in cases when studies report numerous dependent effects for a given outcome type.

### Missing data

The study authors will be contacted if there is missing data in the published articles. When the author cannot be contacted or there is no response from the authors, we will report the study’s characteristics but will exclude it from the meta-analysis.

### Data Analysis and synthesis

Data analysis for this review will be performed using STATA v17. Standardized mean differences (SMDs) for continuous outcome variables and odds ratios (OR) or prevalence ratios (PR) for dichotomous outcome variables will be analyzed separately. Effect sizes will be statistically pooled using inverse variance weighted random effects meta-analysis (29, 30). The random effects model will be used to calculate the pooled mean effect size as the effect size is likely to vary between the different studies (30). Additionally, Random effects models enable statistical inferences to be made to a population of studies other than those included in the meta-analysis (31).

Pooled effects will be expressed using a relevant metric, such as a percentage change in odds or a mean difference measured in natural units of outcome. In studies where multiple effect sizes are reported from the same sample the mean of the combined effect sizes will be calculated. In cases where studies use overlapping samples, an overall estimate will be calculated and those that report effect sizes from independent subgroups, each subgroup will be treated as a separate sample in the meta-analysis.

The synthesis will be presented in the form of a summary of findings tables, simple graphs, and forest plots. This will follow the format of the Cochrane Consumer and Communication Review Group (32). We will describe the included articles, group them based on the study design and type of intervention, organize, and tabulate the results to identify patterns and convert the results into a common descriptive format. These will take the form of outcome data tables, simple graphs, and forest plots, as appropriate. These will be incorporated into the summary of findings tables, which will inform the syntheses for dissemination. We will therefore use both narrative and quantitative synthesis.

### Sensitivity analysis

The sensitivity analysis will be performed by removing studies from the meta-analysis one at a time to determine whether the meta-analysis results are sensitive to any individual study (33). We will also investigate the sensitivity of findings to the level of bias (low risk, some concerns, and high risk).

### Ethics and Dissemination

Ethical approval is not required for this systematic review and meta-analysis as only a secondary analysis of data publicly available in scientific databases will be conducted. The results of this review will be submitted for publication in a peer-reviewed journal. Additionally, the findings will be presented at relevant conferences.

## DISCUSSION

The review will generate findings on the effectiveness of indoor residual spraying for mosquito vector control. The prevalence of malaria in communities following the implementation of the IRS will be reported in this review. Additionally, we shall report on the extent of resistance to WHO pre-qualified insecticides including combination preparations used in IRS for mosquito vector control in sub-Sahara Africa. The prevalence of molecular markers of insecticide resistance among mosquito vectors in SSA will also be reported. We shall also report the factors associated with the effectiveness of indoor residual spraying (IRS) mosquito vector intervention for malaria control in SSA.

This study will systematically collate the evidence available on the effectiveness of WHO pre-qualified insecticides used in indoor residual spraying for malaria mosquito vector control in sub-Saharan Africa. Despite the long-term use of IRS for malaria control, there is limited information on the effectiveness of this vector intervention especially in the presence of other interventions such as LLINs. By collating information about the knockdown effect, susceptibility, residuality (residual efficacy) and moderating factors like types of mosquito vectors, molecular resistance among malaria mosquito vectors, types of insecticides used for IRS and other factors associated with the efficacy of insecticides used in indoor residual spraying in SSA; the findings from this study will provide guidance on the selection of insecticides for use in IRS. This is critical especially due to the current stalling of malaria eradication efforts in most malaria-affected countries.

The effectiveness of mosquito vector control interventions including IRS and LLINs is threatened by the reported emergence and spread of insecticide resistance. This is especially the case as evidenced by the current resurgence of malaria especially in sub-Saharan Africa despite the implementation of multiple vector control interventions. Thus, quality evidence is needed to guide the selection of insecticides to use in mosquito vector control interventions such as IRS and LLINs, especially in high malaria-burdened settings.

### Strengths and limitations of this study

In this review there is no language restriction hence all articles published in all languages will be included if they are eligible. We shall use the GRADE Framework to assess the strength and confidence of commutative evidence. The study evaluates the real-world effectiveness of Deltamethrin-Clothianidin (FLUDORA FUSION) insecticide used in indoor residual spraying in sub-Sahara Africa as an intervention for Malaria control in sub-Saharan Africa hence any findings not in the context of IRS but reporting outcomes related to the use of insecticides will not be considered this will enable pure assessment of the effect of IRS. The review is limited to studies conducted in sub-Saharan Africa and will not account for evidence in studies conducted in other malaria-affected countries

### Amendments

This protocol may be subject to amendments in case of any issues arising in the due course of the review process, should such adjustments be made, they will be reported in the review manuscript and published as deviations from the protocol.

## Data Availability

No datasets were generated or analysed during the current study. All relevant data from this study will be made available upon study completion

## ACKNOWLEDGEMENTS

We would like to thank the team from Infectious Disease Collaboration (IDRC) and Africa Centre for Systematic Reviews and Knowledge Translation, Makerere University College of Health Sciences for their assistance in critically reviewing the content of the protocol.

## AUTHOR CONTRIBUTIONS

Conceptualization of the study was done by (MO, NL, KOO). MO, NL, SN, EA, HM and KOO drafted the protocol, Critical review (MO, NL, KOO, AAK, RA, KG, SN, EA & HM), and approval of the final version (MO, NL, KOO, AAK, RA, KG, SN, EA & HM).

## SUPPORTING INFORMATION

**S1:** PRISMA_P checklist

## REFERENCES

1. World Health Organization. Indoor residual spraying: an operational manual for indoor residual spraying (IRS) for malaria transmission control and elimination: World Health Organization; 2015.

2. Organization. WH. Global Malaria Programme. Global plan for insecticide resistance management in malaria vectors. Geneva: World Health Organization; 2012.

3. Tangena J-AA, Hendriks CM, Devine M, Tammaro M, Trett AE, Williams I, et al. Indoor residual spraying for malaria control in sub-Saharan Africa 1997 to 2017: an adjusted retrospective analysis. Malaria journal. 2020;19(1):1–15.

4. Mint Mohamed Lemine A, Ould Lemrabott MA, Niang EHA, Basco LK, Bogreau H, Faye O, et al. Pyrethroid resistance in the major malaria vector Anopheles arabiensis in Nouakchott, Mauritania. Parasites & vectors. 2018;11(1):1-8.

5. Abeku TA, Helinski ME, Kirby MJ, Ssekitooleko J, Bass C, Kyomuhangi I, et al. Insecticide resistance patterns in Uganda and the effect of indoor residual spraying with bendiocarb on kdr L1014S frequencies in Anopheles gambiae ss. Malaria Journal. 2017;16:1–11.

6. Epstein A, Maiteki-Sebuguzi C, Namuganga JF, Nankabirwa JI, Gonahasa S, Opigo J, et al. Resurgence of malaria in Uganda despite sustained indoor residual spraying and repeated long lasting insecticidal net distributions. PLOS Global Public Health. 2022;2(9):e0000676.

7. Coleman M, Hemingway J, Gleave KA, Wiebe A, Gething PW, Moyes CL. Developing global maps of insecticide resistance risk to improve vector control. Malaria journal. 2017;16:1–9.

8. Organization WH. Global plan for insecticide resistance management in malaria vectors: World Health Organization; 2012.

9. Syme T, Fongnikin A, Todjinou D, Govoetchan R, Gbegbo M, Rowland M, et al. Which indoor residual spraying insecticide best complements standard pyrethroid long-lasting insecticidal nets for improved control of pyrethroid resistant malaria vectors? PloS one. 2021;16(1):e0245804.

10. Fongnikin A, Houeto N, Agbevo A, Odjo A, Syme T, N’Guessan R, et al. Efficacy of Fludora® Fusion (a mixture of deltamethrin and clothianidin) for indoor residual spraying against pyrethroid-resistant malaria vectors: laboratory and experimental hut evaluation. Parasites & Vectors. 2020;13:1–11.

11. Kafy HT, Ismail BA, Mnzava AP, Lines J, Abdin MSE, Eltaher JS, et al. Impact of insecticide resistance in Anopheles arabiensis on malaria incidence and prevalence in Sudan and the costs of mitigation. Proceedings of the National Academy of Sciences. 2017;114(52):E11267–E75.

12. Zhou Y, Zhang W-X, Tembo E, Xie M-Z, Zhang S-S, Wang X-R, et al. Effectiveness of indoor residual spraying on malaria control: a systematic review and meta-analysis. Infectious Diseases of Poverty. 2022;11(04):29–42.

13. Gimnig JE, Steinhardt LC, Awolola TS, Impoinvil D, Zohdy S, Lindblade KA. Reducing Malaria Transmission through Reactive Indoor Residual Spraying: A Systematic Review. The American Journal of Tropical Medicine and Hygiene. 2023:tpmd220745-tpmd.

14. Pryce J, Medley N, Choi L. Indoor residual spraying for preventing malaria in communities using insecticide-treated nets. Cochrane Database of Systematic Reviews. 2022(1).

15. Page MA-O, Moher D, Bossuyt PM, Boutron I, Hoffmann TC, Mulrow CD, et al. PRISMA 2020 explanation and elaboration: updated guidance and exemplars for reporting systematic reviews. 2021(1756-1833 (Electronic)).

16. Moher D, Shamseer L, Clarke M, Ghersi D, Liberati A, Petticrew M, et al. Preferred reporting items for systematic review and meta-analysis protocols (PRISMA-P) 2015 statement. Systematic Reviews. 2015;4(1):1.

17. Wells G. The Newcastle-Ottawa Scale (NOS) for assessing the quality of nonrandomised studies in meta-analysis. 2004.

18. Higgins JP, Altman DG, Gøtzsche PC, Jüni P, Moher D, Oxman AD, et al. The Cochrane Collaboration’s tool for assessing risk of bias in randomised trials. Bmj. 2011;343.

19. Sterne JA, Hernán MA, Reeves BC, Savović J, Berkman ND, Viswanathan M, et al. ROBINS-I: a tool for assessing risk of bias in non-randomised studies of interventions. bmj. 2016;355.

20. Thomson H, Craig P, Hilton-Boon M, Campbell M, Katikireddi SV. Applying the ROBINS-I tool to natural experiments: an example from public health. Systematic reviews. 2018;7(1):1–12.

21. Duval S, Tweedie R. Trim and fill: A simple funnel-plot-based method of testing and adjusting for publication bias in meta-analysis. 2000(0006-341X (Print)).

22. Egger M, Davey Smith G Fau - Schneider M, Schneider M Fau - Minder C, Minder C. Bias in meta-analysis detected by a simple, graphical test. 1997(0959-8138 (Print)).

23. Schünemann HJ, Cuello C, Akl EA, Mustafa RA, Meerpohl JJ, Thayer K, et al. GRADE guidelines: 18. How ROBINS-I and other tools to assess risk of bias in nonrandomized studies should be used to rate the certainty of a body of evidence. 2013(1878-5921 (Electronic)).

24. Higgins JP. Commentary: Heterogeneity in meta-analysis should be expected and appropriately quantified. 2009(1464-3685 (Electronic)).

25. Higgins JP, Thompson SG. Quantifying heterogeneity in a meta-analysis. Statistics in medicine. 2002;21(11):1539–58.

26. Borenstein M, Cooper H, Hedges L, Valentine J. Heterogeneity in meta-analysis. The handbook of research synthesis and meta-analysis. 2019;3:453–70.

27. Huedo-Medina TB, Sánchez-Meca J, Marín-Martínez F, Botella J. Assessing heterogeneity in meta-analysis: Q statistic or I² index? Psychological methods. 2006;11(2):193.

28. López-López JA-O, Page MJ, Lipsey MW, Higgins JPT. Dealing with effect size multiplicity in systematic reviews and meta-analyses. LID - 10.1002/jrsm.1310 [doi]. (1759-2887 (Electronic)).

29. Hunter JE, Schmidt FL. Methods of meta-analysis: Correcting error and bias in research findings: Sage; 2004.

30. Gegenfurtner A. Comparing Two Handbooks of Meta-Analysis: Review of Hunter & Schmidt, Methods of Meta-Analysis: Correcting Error and Bias in Research Findings, and Borenstein, Hedges, Higgins, and Rothstein, Introduction to Meta-Analysis. Springer; 2011.

31. Basu A. An Introduction to Meta-Analysis. A Guide to the Scientific Career: Virtues, Communication, Research and Academic Writing. 2019:615–38.

32. Ryan R. Cochrane Consumers and Communication Review Group. ‘Cochrane Consumers and Communication Review Group: data synthesis and analysis’. 2013.

33. Copas J, Shi JQ. Meta-analysis, funnel plots and sensitivity analysis. Biostatistics. 2000;1(3):247–62.

